# Characteristics and Outcomes of Mavacamten Use In 2440 Patients with Obstructive Hypertrophic Cardiomyopathy

**DOI:** 10.1101/2025.03.24.25324576

**Authors:** Ingy Mahana, Angelica T. Mejia, Miriam R. Elman, Daniel Kamna, Olives Nguyen, Hailey Volk, Hongya Chen, Ahmad Masri

**Author notes:** Corresponding author: Ahmad Masri, MD MS, Assistant Professor of Medicine, Mail code: UHN-62, 3181 SW Sam Jackson Rd Portland, OR 97239.

## Abstract

**Aims:** To assess real-world characteristics, outcomes, and healthcare utilization associated with commercial mavacamten use in patients with oHCM.

**Methods and Results:** We used the Symphony Dataverse to study patients receiving mavacamten between May 25, 2022 and June 30, 2024. Safety outcomes were prespecified and individual hospital-level claims manually reviewed to categorize cardiovascular hospitalizations. We included 2,440 patients and 18,494 mavacamten dispenses (63% females, median age 66 years [Interquartile range (IQR): 55, 73]). Patients were followed for a median of 3,037.0 days (IQR: 2,808, 3,221) prior to mavacamten initiation, and for 240 days (IQR: 117, 407) on mavacamten. Atrial fibrillation or flutter (AF/AFL) at baseline were present in 536 (22%) of patients, while new-onset AF/AFL requiring therapy occurred in 125 patients (5%) and new heart failure (HF) in 100 patients (4%). During follow-up, 154 patients (6%) received new dispenses of loop diuretics, 102 (4%) oral anticoagulants, 78 (3%) amiodarone, 17 (0.7%) sotalol, and 6 (0.2%) dofetilide. There was a total of 306 patients (13%) with 428 acute care episodes (most common claims were HF (n= 80, 26%), AF (n=74, 24%), and AFL (n= 16, 5%)). Predictors of safety outcomes (new HF, new AF/AFL, ventricular tachycardia, or transient ischemic attack [TIA]/stroke claim) were: age at mavacamten start, obesity, and TIA/stroke history.

**Conclusion:** With 8 month median follow-up in this study, healthcare utilization for patients on mavacamten appeared substantial. New AF/AFL and HF occurred in 5% and 4%, respectively. Further work is needed to understand longer-term outcomes of mavacamten use in oHCM.

## Introduction

Hypertrophic cardiomyopathy (HCM) is common, and obstructive HCM (oHCM) constitutes two-thirds of patients. The underlying mechanism of dynamic left ventricular outflow tract (LVOT) obstruction is explained by hypercontractility, which leads to progressive septal hypertrophy, septal contact, systolic anterior motion of the mitral valve causing dynamic left ventricular outflow tract obstruction^1,2^. This obstruction causes an increase in left ventricular pressure and impairs diastolic function, which eventually decreases cardiac output and causes heart failure. Patients’ symptomatology varies from chest pain, shortness of breath, palpitations from underlying arrhythmias, and syncope. For long, oHCM conventional medical treatment using beta blockers, non-dihydropyridine calcium channel blockers, and disopyramide was the mainstay of medical treatment until the first cardiac myosin inhibitor, mavacamten was approved by the United States Food and Drug Administration in 2022 and the European Medicines Agency in 2023. Mavacamten is currently indicated as a second-line therapy for treating patients with symptomatic oHCM to improve symptoms and functional capacity^3,4^.

Mavacamten underwent multiple evaluations in clinical trials. In Explorer-HCM, mavacamten improved peak oxygen consumption and New York Heart Association (NYHA) class as compared to placebo^5^. However, mavacamten use over 30 weeks caused an excessive reduction of left ventricular ejection fraction (LVEF) to <50% in 5.7% of patients. A second phase III trial, VALOR-HCM, enrolled patients who were referred to septal reduction therapy, and showed that over 112-128 weeks, 15 (13.8%) patients had developed LVEF<50%, including 2 with LVEF<30%, and 1 death occurred in a close proximity to the low LVEF event. The United States Food and Drug Administration (FDA) approved mavacamten based on the EXPLORER-HCM trial results, and due to the decision to not use mavacamten dose in monitoring patients, a rigorous and restrictive risk mitigation program (REMS) was instituted^6^. Since the start of commercial use of mavacamten, there have been multiple small single- or multi-center studies assessing the efficacy and safety of mavacamten. However, these studies enrolled small numbers and occurred in experienced centers. In addition, atrial fibrillation and flutter (AF/AFL) are emerging as a frequently encountered issue in patients with oHCM on mavacamten^7–9^. In this study, we leveraged a national all-payers medical and pharmacy claims database to characterize the outcomes of mavacamten use in a real-world cohort.

## Methods

This population-based, observational study used claims data from the Symphony Heath Integrated Dataverse from January 1, 2015 – June 30, 2024 to construct a cohort of patients dispensed mavacamten following its approval in the United States. The Integrated Dataverse is a nationally representative, provider-based claims database containing longitudinal patient medical data for claims submitted to all payer types (commercial, cash, Medicare/Medicaid, etc.). We included patients between 12 and 80 years of age with at least one claim for mavacamten between May 25, 2022 and June 30, 2024. Patients older than 80 years are still coded as 80 years.

The study’s start date was the first claim for mavacamten in the database. Baseline clinical characteristics were assessed at any point prior to the index (start) date except medication history, obesity, tobacco use, hyperlipidemia, which were identified in the 90 days prior to starting mavacamten. Diagnoses, procedures/surgeries, and healthcare utilization was defined using International Classification of Diseases, 10th revision (ICD-10); Current Procedural Terminology (CPT), and ICD-10 Procedural Coding System (ICD-10-PCS) codes (Table S1, Online Supplement). Information on mavacamten including fill date, days supply, and strength were obtained from fills as were dispenses for other medications/medication classes (Table S2, Online Supplement). Follow-up was defined as the period between the index date and end of clinical activity, based on patients’ last claim in the Symphony database. Episodes of mavacamten use were characterized as consecutive fills with a gap of 35 days or less, using days’ supply to calculate the end date for mavacamten coverage on each claim. Treatment discontinuation was designated as a period of ≥90 days from the end of patients’ first mavacamten episode to the end of clinical activity in the database.

We assessed the characteristics of mavacamten fills as well as patient diagnoses, procedures, and medications in the follow-up period. For the subset of our cohort with acute care encounters, we manually reviewed diagnosis, procedural, surgical, and medication claims within 14 days of the event to determine potentially related conditions and classify them. In addition to descriptive outcomes, we defined two composite outcomes: (1) cardiovascular diagnoses related to mavacamten use including new HF, new AF, new AFL, ventricular tachycardia (VT), or transient ischemic attack/stroke diagnosis (“cardiovascular diagnoses”) and (2) cardiovascular diagnoses related to mavacamten in (1) as well as new use of loop diuretics, which is used as a surrogate for symptoms of volume overload in patients with HCM. Time from mavacamten initiation to the first of the composite events or end of clinical activity was computed for each composite outcome.

### Statistical Analysis

Patient demographics and clinical characteristics were summarized using frequency with percentage for categorical variables and median with interquartile range (IQR) for continuous variables. Cumulative incidence of first discontinuation of mavacamten and composite events were visualized with Kaplan-Meier plots. Cox proportional hazards models were used to identify characteristics associated with each composite outcome. The following candidate variables were selected *a priori* based on clinical knowledge: patient age at mavacamten initiation; sex; United States region (West, Midwest, South, and Northeast); weeks present in symphony database prior to mavacamten initiation; baseline comorbidities including obesity, hypertension, diabetes, stroke, coronary artery disease, myocardial infarction, chronic kidney disease, sleep apnea; and beta-blockers and calcium channel blockers use before mavacamten. Variable selection was performed on the set of candidate variable using backwards elimination using the Akaike information criterion (AIC), equivalent to a p<0.157 stopping criterion^10^. Hazard ratios (HR) from univariable, global, and backwards elimination models are reported with 95% (CI) confidence intervals.

Model stability investigations were conducted to assess model and variable uncertainty; additional details and results of these stability analyses are included in the Online Supplement (Tables S4-7)^10,11^. SAS v9.4 (SAS Institute, Cary, NC) was used for data management and R 4.3.1 (R Core Team: R: A language and environment for statistical computing, Vienna, Austria, R Foundation for Statistical Computing, 2019) for analyses.

## Results

From May 2022 to June 2024, 2440 patients received mavacamten and were included in the study cohort. The median age was 66.0 (Interquartile range [IQR]: 56.0, 73.0) years and 1,530 (62.7%) were female. Patients were enrolled and followed for a median of 3,037.0 (IQR: 2,808.0, 3,220.5) days prior to being prescribed mavacamten. Notable baseline comorbidities included hypertension in 1,843 patients (75.5%), diabetes in 644 patients (26.4%), coronary artery disease in 969 patients (39.7%), and obesity in 242 patients (9.9%). History of claims reflecting AF/AFL were common, seen in 715 patients (29.3%) (Table 1). The majority of patients were on background medical therapy including 1,326 (54.3%) on beta-blockers, 689 (28.2%) on calcium channel blockers, and 129 (5.3%) on disopyramide).

**Table 1.** Baseline characteristics of patients initiating mavacamten^a^.

|  |  |
| --- | --- |
| **Characteristic** | **Overall Cohort**<br>**(n=2,440)** |
| Female | 1,530 (62.7) |
| Age at first mavacamten fill, years, median (IQR) [range] | 66.0 (56.0, 73.0) [12.0, 80.0] |
| United States region |  |
| West | 430 (17.6) |
| Midwest | 513 (21.0) |
| South | 901 (36.9) |
| Northeast | 596 (24.4) |
| Comorbidity history |  |
| Obesity | 242 (9.9) |
| Tobacco use | 84 (3.4) |
| Hypertension | 1,843 (75.5) |
| Hyperlipidemia | 492 (20.2) |
| Diabetes | 644 (26.4) |
| Peripheral artery disease | 265 (10.9) |
| Peripheral vascular disease | 186 (7.6) |
| Stroke/TIA | 253 (10.4) |
| Coronary artery disease | 969 (39.7) |
| Myocardial infarction | 366 (15.0) |
| Chronic kidney disease | 386 (15.8) |
| Sleep apnea | 805 (33.0) |
| COPD | 186 (7.6) |
| Indication of AF/FL<sup>b</sup> | 715 (29.3) |
| Procedure/Surgical history |  |
| CABG | 54 (2.2) |
| Heart valve surgery | 65 (2.7) |
| PCI | 24 (1.0) |
| Prescription history |  |
| Beta blocker | 1,326 (54.3) |
| Calcium channel blocker | 689 (28.2) |
| Disopyramide | 129 (5.3) |
| Days from first symphony claim to first mavacamten fill, median (IQR) [range] | 3,037.0 (2,808.0, 3,220.5) [0.0, 3,465.0] |
| Days from first mavacamten fill to last symphony claim, median (IQR) [range] | 240.0 (117.0, 407.0) [0.0, 762.0] |
| Abbreviations: IQR=interquartile range, TIA=transient ischemic attack, COPD=chronic obstructive pulmonary disease, AF/FL=atrial fibrillation/atrial flutter, CABG=coronary artery bypass graft, PCI=percutaneous coronary intervention. |  |
| <sup>a</sup>Data are n (%) except where noted; |  |
| <sup>b</sup>AF/FL definition based on diagnosis, procedure/surgery, or medication-based indication. |  |

### Follow-up and outcomes

Patients were followed for a median of 240.0 (IQR: 117.0, 407.0) days following start of mavacamten. The median dose of mavacamten was 5.0 (IQR: 5.0, 10.0) mg with a median number of fills of 6.0 (IQR: 2.0, 12.0). During the follow-up period, 1,075 (44.1%) patients had dose modification, with the majority being down-titrations as detailed in Table S3 in the online supplement. A total of 18,494 fills were captured during the study period. During the follow-up period, chest pain and angina were common, and occurred in 191 (7.8%) patients. New stress-induced cardiomyopathy was rare, with claims in only 2 patients (0.1%). In addition, new AF/AFL was common and occurred in 125 patients (5.1%), while new heart failure occurred in 99 patients (4.1%). Other diagnoses reported during follow-up are presented in Table 2 as are new medications of interest. There were 154 patients (6.0%) who received new dispenses of loop diuretics, while 102 (4.2%) received oral anticoagulants, 78 (3.2%) received amiodarone, 17 (0.7%) received sotalol, and 6 (0.2%) received dofetilide. Over the follow-up period, there was a total of 306 patients (12.5%) with 428 acute care claims. Heart failure was the most common reason, occurring in 80 patients (26.1% of claims, 3.3% of all patients) after a median of 108.5 days (IQR: 35.0, 225.0) from mavacamten initiation. Syncope necessitating acute care occurred in 28 patients (9.2% of claims, 1.2% of all patients). Arrhythmias were another common cause of acute care encounters. There were 74 patients (24.2% of claims, 3.0% of all patients) who had AF at a median of 95.5 days (53.0, 191.0), 16 (5.2% of claims, 0.7% of all patients) with atrial flutter leading to acute care, and 23 (7.5% of claims, 0.9% of all patients) with stroke. Acute care encounters also included encounters for atrial arrhythmia management with cardioversion or ablation, and a few patients transitioned to myectomy. Table 3 summarizes the diagnoses associated with acute care encounters and the median duration from mavacamten exposure.

**Table 2.** Diagnoses, prescriptions, and healthcare utilization during follow-up after mavacamten initiation^a^.

| Characteristic | Overall cohort<br>(n=2,440) |
| --- | --- |
| Diagnoses |  |
| Angina | 21 (0.9) |
| Dyspnea | 287 (11.8) |
| Chest pain | 178 (7.3) |
| Fatigue | 84 (3.4) |
| Palpitations | 115 (4.7) |
| Edema | 47 (1.9) |
| Syncope | 72 (3.0) |
| New stress cardiomyopathy | 2 (0.1) |
| Any indication of AF/FL<sup>b</sup> | 536 (22.0) |
| New indication of AF/FL<sup>b</sup> | 125 (5.1) |
| New ventricular tachycardia | 26 (1.1) |
| New heart failure | 99 (4.1) |
| New depression | 38 (1.6) |
| Acute care encounter |  |
| Hospitalization/observation | 165 (6.8) |
| Emergency department | 230 (9.4) |
| Critical care | 33 (1.4) |
| New prescriptions |  |
| Oral anticoagulant | 102 (4.2) |
| Thiazide diuretic | 56 (2.3) |
| Potassium sparing diuretic | 87 (3.6) |
| Loop diuretic | 154 (6.3) |
| Sotalol | 17 (0.7) |
| Amiodarone | 78 (3.2) |
| Dofetilide | 6 (0.2) |

Abbreviations: AF/FL=atrial fibrillation/atrial flutter.
<sup>a</sup>Data are n (%);
<sup>b</sup>AF/FL definition based on diagnosis, procedure/surgery, or medication-based indication.

**Table 3.**
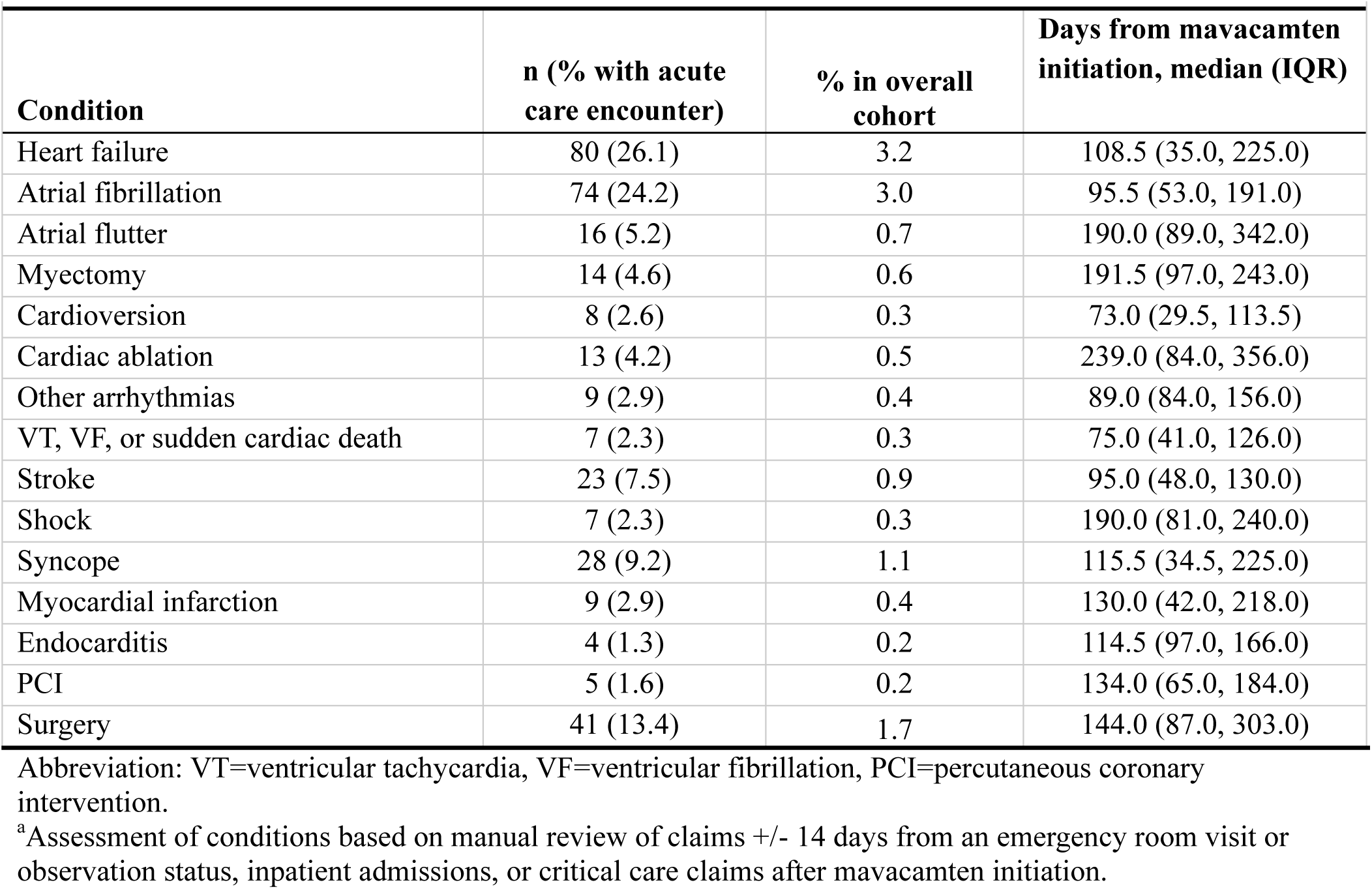
Diagnoses in 306 patients with claims for acute care encounters after mavacamten initiation^a^.

### Discontinuations

Out of 2,440 patients, 412 (16.9%) had a discontinuation after their first mavacamten episode without an additional mavacamten fill in ≥90 days. An additional 85 (3.5%) individuals had a ≥90 day interruption after an initial episode but restarted mavacamten for at least one additional claim. For those who had acute care encounter, 24 (7.8% of 306) discontinued mavacamten, and 22 (91.7%) of them did not appear to have restated mavacamten.

### Predictors of adverse outcomes on mavacamten

In the model with composite cardiovascular diagnoses, time to the composite event was associated with age at first mavacamten fill (adjusted HR [aHR]: 1.01; 95% CI: 1.00, 1.02; p=0.0066) and the comorbidities obesity (aHR: 1.56; 95% CI: 1.14, 2.14; p=0.0052) and stroke/TIA (aHR: 1.97; 95% CI: 1.47, 2.64; P<0.0001) were associated with the composite outcome of an adverse event of interest (Figure 1A and Table 4). The inclusion of loop diuretic in the second model resulted in a slight change in the predictors of the composite outcome to include beta blocker use (aHR 1.23; 95% CI: 1.01, 1.49; p=0.0397) and myocardial infarction (aHR: 1.29; 95% CI 1.01, 1.65; p=0.0425) along with age (aHR: 1.02, 95% CI: 1.01, 1.02; p<0.0001), obesity (aHR: 1.39; 95% CI: 1.05, 1.85; p=0.0208), and stroke/TIA (aHR: 1.75; 95% CI: 1.35, 2.28; p<0.0001) (Figure 1B and Table 5).

**Figure 1.**
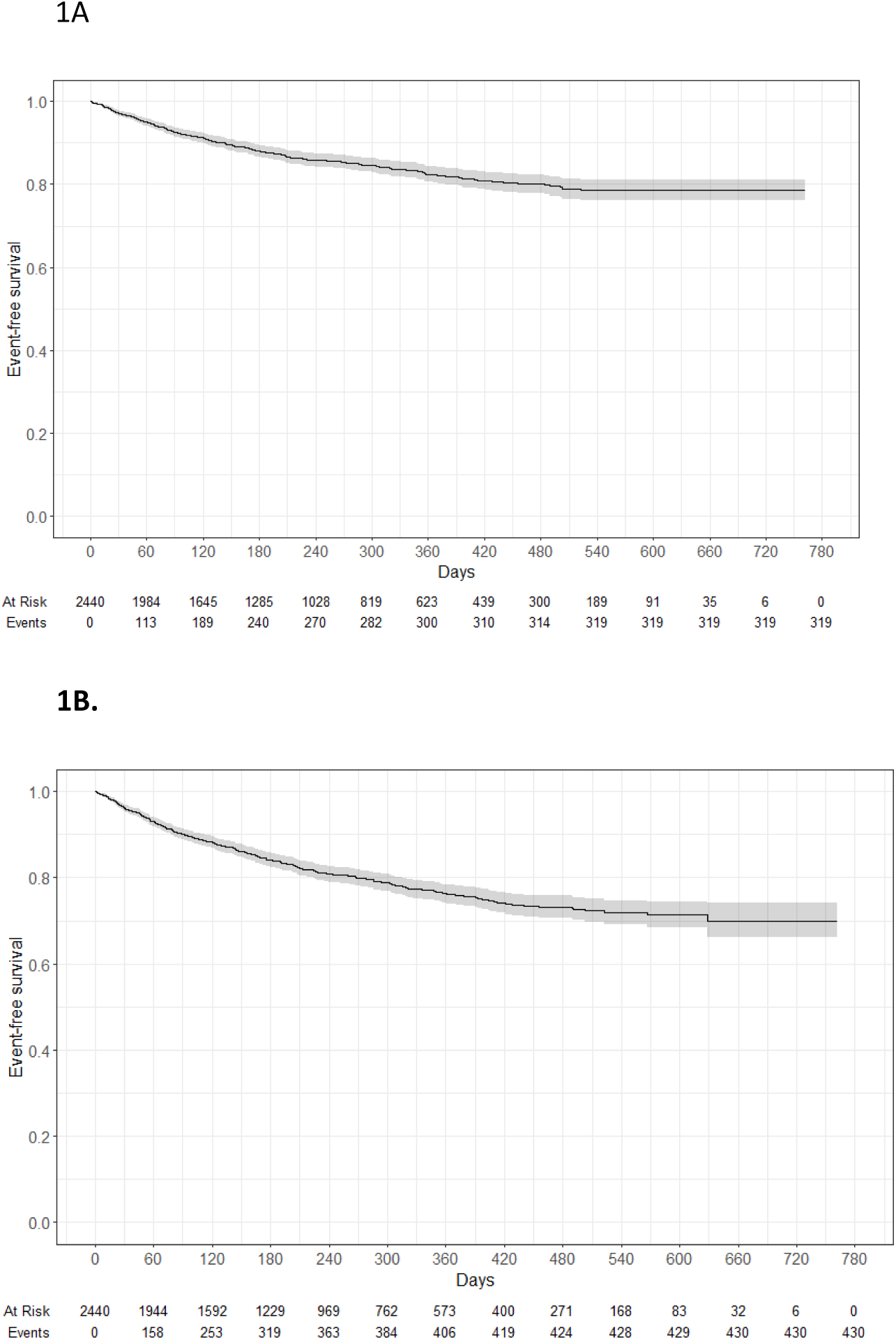
Outcomes in patients receiving mavacamten, IA) Kaplan-Meier curve for cumulative incidence of composite event for cardiovascular diagnoses related to mavacamten use and IB) Kaplan-Meier curve for cumulative incidence of composite event for cardiovascular diagnoses related to mavacamten use or new loop diuretic

**Table 4.**
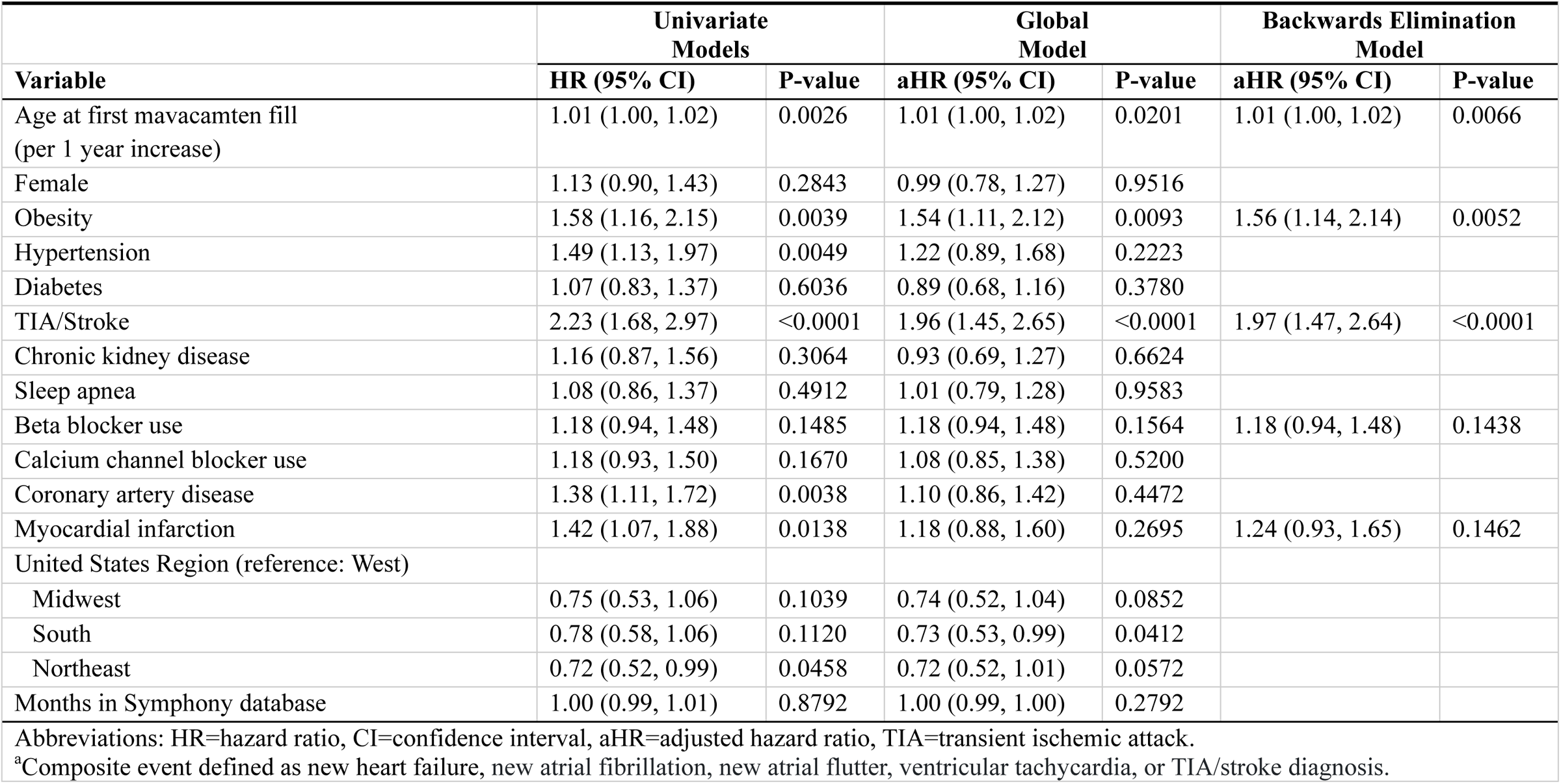
Model estimates for baseline variables associated with composite outcome based on cardiovascular diagnoses^a^.

**Table 5.** Model estimates for baseline variables associated with composite outcome based on cardiovascular diagnoses and new loop diuretic claim^a^.

|  | **Univariate Models** |  | **Global Model** |  | **Backwards Elimination Model** |  |
| --- | --- | --- | --- | --- | --- | --- |
| **Variable** | **HR (95% CI)** | **P-value** | **aHR (95% CI)** | **P-value** | **aHR (95% CI)** | **P-value** |
| Age at first mavacamten fill (per 1 year increase) | 1.02 (1.01, 1.02) | <0.0001 | 1.01 (1.01, 1.02) | 0.0015 | 1.02 (1.01, 1.02) | <0.0001 |
| Female | 1.26 (1.03, 1.55) | 0.0223 | 1.10 (0.89, 1.36) | 0.3724 |  |  |
| Obesity | 1.39 (1.05, 1.84) | 0.0195 | 1.36 (1.02, 1.82) | 0.0370 | 1.39 (1.05, 1.85) | 0.0208 |
| Hypertension | 1.34 (1.06, 1.69) | 0.0147 | 1.07 (0.82, 1.40) | 0.6223 |  |  |
| Diabetes | 1.17 (0.95, 1.44) | 0.1465 | 1.00 (0.80, 1.26) | 0.9711 |  |  |
| TIA/Stroke | 2.02 (1.56, 2.61) | <0.0001 | 1.73 (1.32, 2.27) | <0.0001 | 1.75 (1.35, 2.28) | <0.0001 |
| Chronic kidney disease | 1.23 (0.96, 1.58) | 0.0953 | 1.00 (0.77, 1.30) | 0.9787 |  |  |
| Sleep apnea | 1.12 (0.92, 1.36) | 0.2733 | 1.06 (0.86, 1.31) | 0.5758 |  |  |
| Beta blocker use | 1.22 (1.00, 1.48) | 0.0462 | 1.24 (1.01, 1.50) | 0.0350 | 1.23 (1.01, 1.49) | 0.0397 |
| Calcium channel blocker use | 1.11 (0.91, 1.37) | 0.3119 | 1.03 (0.83, 1.26) | 0.8042 |  |  |
| Coronary artery disease | 1.27 (1.05, 1.53) | 0.0152 | 0.99 (0.80, 1.24) | 0.9493 |  |  |
| Myocardial infarction | 1.43 (1.12, 1.82) | 0.0038 | 1.26 (0.97, 1.63) | 0.0819 | 1.29 (1.01, 1.65) | 0.0425 |
| United States region (reference: West) |  |  |  |  |  |  |
| Midwest | 0.94 (0.70, 1.26) | 0.6586 | 0.92 (0.68, 1.24) | 0.5795 |  |  |
| South | 0.86 (0.66, 1.12) | 0.2536 | 0.80 (0.61, 1.06) | 0.1170 |  |  |
| Northeast | 0.77 (0.58, 1.04) | 0.0850 | 0.79 (0.59, 1.06) | 0.1142 |  |  |
| Months in Symphony database | 1.00 (1.00, 1.01) | 0.6998 | 1.00 (0.99, 1.00) | 0.4372 |  |  |

Abbreviations: HR=hazard ratio, CI=confidence interval, aHR=adjusted hazard ratio, TIA=transient ischemic attack
<sup>a</sup>Composite event defined as new loop diuretic claim on new heart failure, new atrial fibrillation, new atrial flutter, ventricular tachycardia, or TIA/stroke diagnosis.

## Discussion

Mavacamten is the first-in-class CMI. It revolutionized the treatment of patients with oHCM. Safety and efficacy of mavacamten were derived from rigorously conducted clinical trials, which usually included core-lab confirmation of echocardiographic diagnosis of oHCM. However, since commercial use is not subjected to the same rigorous patient selection and monitoring, there is a need to understand the safety of mavacamten in a large, unselected patient cohort. In this current study, patients with oHCM treated with mavacamten had frequent acute care encounters, with the majority related to heart failure and/or atrial arrhythmias. Patients had significant utilization of additional pharmacotherapies including oral anticoagulation, anti-arrhythmic drugs, and diuretics. In addition, a significant number of patients discontinued mavacamten during follow-up even when a stringent 90-day interruption period was used.

Mavacamten was approved by the FDA in 2022 and by the EMA in 2023. Due to its potential to cause systolic dysfunction and heart failure, it was required to have a REMS program in the United States or metabolizer status genotyping and monitoring in Europe^3,4^. The REMS monitoring program is onerous and uses echocardiography as a surrogate to mavacamten drug-level, since the drug level was used in the pivotal phase III EXPLORER-HCM trial, but was not required by the FDA for commercial use. In addition, drug-drug interactions due to mavacamten metabolism pathways require dispensing pharmacies to review patient’s medications and supplements at each dispense. In Europe, a distinct monitoring program is recommended which also includes CYP genotyping.

The instituted REMS monitoring program is distinct from how the clinical trials were run, and as such, commercial use of mavacamten requires further scrutiny. Despite almost 3 years since the approval of mavacamten in the United States, there is a paucity of large-scale evaluations of patient outcomes. The single-center studies are informative, but they suffer from small sample size, and selection and reporting biases^12–15^. In 2023 a real-world short-term retrospective cohort study of 659 patients followed for 118 days showed high adherence to mavacamten^16^. In addition, the severe adverse events from mavacamten are expected to be uncommon, therefore, a large representative sample is required.

Recent data published from the FDA REMS program showed a 6.1% discontinuation rate of mavacamten after the first fill, 4.6% of patients had LVEF <50%, 1.3% had heart failure hospitalization, and 0.3% had both LVEF <50% and heart failure hospitalization. However, these data do not account for patients who discontinued mavacamten permanently due an adverse event. As such, there inherent survival and reporting biases to such data since there is no obligation to report on patients who simply discontinued mavacamten without plans to resume and therefore the need to fulfill the REMS program requirement^8^. A better suited database for this question is one that allows for understanding of the events that occur to patients who permanently discontinued or temporarily held mavacamten.

Mavacamten has already been adopted by the American College of Cardiology/American Heart Association guidelines as a second-line therapy for symptomatic oHCM, adjacent to disopyramide and septal reduction therapy (SRT), while the European Society of Cardiology guidelines recommend it as a second-line therapy prior to proceeding with SRT^3,4^. The EXPLORER-HCM and VALOR-HCM trials have shown the ability of mavacamten to improve exercise capacity, symptoms, and avert the need for SRT^(8,18)^. However, only VALOR-HCM trial based dose titration solely on echocardiographic monitoring, which resembles real-world practice. In VALOR-HCM, over 112-128 weeks of mavacamten treatment, 15 (13.8%) patients developed LVEF <50%^9^.

In the current study, our goal was not to re-define the incidence of LVEF <50% on mavacamten. We sought to study healthcare utilization in the acute care settings as well as new diagnoses of interest and discontinuation rates. Another goal is to use pharmacy claims to study disease progression and adverse events. In HCM, symptoms suggestive of heart failure directly overlap with symptoms related to HCM; therefore, other tools are required to study disease evolution in patients. An important surrogate of patients’ well-being in HCM is the new use of loop diuretics, which signals the transition to a more complex disease state. While loop diuretics use should not equate heart failure or progressive end-stage HCM, it is a reasonable surrogate of how the disease is evolving. In our study, loop diuretics were newly introduced in 154 patients (6.0%) after mavacamten had been initiated.

Aside from heart failure, AF and AFL, and their respective treatments, are emerging as a significant contributor to morbidity while on mavacamten treatment, and beyond. Developing new-onset AF/AFL mandates lifelong anticoagulation in patients with HCM irrespective of other risk factors, and as such, the morbidity associated with this diagnosis carries over a lifetime. AF/AFL are commonly encountered in HCM, with a prevalence as high as 20-30%, and a typical incidence of 2% per year^17–20^. It remains unclear if there is an association between mavacamten, or cardiac myosin inhibition, and AF/AFL. However, there are some clues that the incidence of new-onset AF/AFL might be higher than expected, especially with how effective mavacamten is in lowering LVOT gradients and improving left atrial volume^21^. In a study of human atrial tissue, mavacamten inhibited the α-myosin heavy chain in the atria at a similar rate to the β-myosin heavy chain found in the ventricle, with resultant reduction in atrial twitch amplitude and possibly depressing atrial contractility^22^. Whether these findings translate into humans with HCM remains to be seen.

Data form clinical trials provide further insights. AF and AFL occurred in 7.7% and 7.1% of patients on mavacamten compared to 6.5% and 0% on placebo in the EXPLORER-HCM and VALOR-HCM trials, respectively. The long term extension study MAVA-LTE showed 33 (14.3%) events of AF/AFL with 18 (7.8%) being new-onset over 166 weeks of follow up, and the long-term extension study of VALOR-HCM showed new-onset AF/AFL in 11 patients (10.2%) over 128 weeks^7,9^. In the current study, new AF/AFL was common and occurred in 125 patients (5.1%), leading to a 7.6% annualized rate of AF/AFL in the first year. The majority of new-onset AF/AFL episodes required a rhythm control strategy with an initiation of oral anticoagulation. Combined with prior findings from mavacamten trials, these findings represent an urgent need for an in-depth study of this phenomenon. Further data are needed to understand if AF/AFL episodes are related to duration of mavacamten exposure, metabolizer status, hemodynamic change, effect on atrial myosin, or if these are simply representations of a common arrhythmia that occurs in patients with oHCM.

Finally, discontinuations of mavacamten were substantial. We chose to focus on 90 days or more in order to have a more stringent and specific criteria. Using this cut-off, 412 (16.9%) patients had discontinued mavacamten after their first episode without an additional fill, while an additional 85 (3.5%) individuals had more than 90-day interruption after an initial episode but restarted mavacamten for at least one additional fill. These longer-term discontinuations likely signal permanent discontinuation of mavacamten. While temporary hold of mavacamten typically lasts 4-6 weeks for LVEF<50%, we felt that it is rather difficult to discern these short-term temporary holds.

Our study has multiple unavoidable limitations. First, using claims data provided access to a large representative sample across the country at the expense of granular phenotypic, laboratory and echocardiographic data. The goal of this analysis is to quantify patterns of mavacamten use, complications, and acute care utilization. Despite almost 3 years of commercial availability of mavacamten, no large-scale real-world studies have been conducted. Secondly, while the sample size is large enough for a disease like HCM on a specialty drug such as mavacamten, this database does not capture all patients in the United States who received mavacamten. Thirdly, we are unable to assess mortality in this database. The Symphony database was specifically used to access details of pharmacy claims that provided us with significant insights into complications and morbidity while on mavacamten using fill claims of new medications. In addition, we manually reviewed all the acute care claims codes and timelines, classifying them into the categories in Table 3. Finally, we did not include a control group due to the inherent heterogeneity of patients who are selected to receive mavacamten vs other traditional therapies oHCM.

## Conclusion

Over a median of 8 months, healthcare utilization and claims for patients on mavacamten appeared substantial. New AF/AFL and heart failure occurred frequently and required treatment with loop diuretics, rhythm control, and anticoagulation in the majority. Patients on mavacamten had frequent acute care encounters, and rates of discontinuation were substantial. Further work is needed to understand the longer-term outcomes of mavacamten use in oHCM, and whether these findings represent the natural history of oHCM or if they represent particular complications of mavacamten use.

## Data Availability

Data are available through Symphony and authors are not allowed to share the data directly.

## Acknowledgement

none

## References

1. Marian AJ, Braunwald E. Hypertrophic Cardiomyopathy: Genetics, Pathogenesis, Clinical Manifestations, Diagnosis, and Therapy. Circ Res. 2017;121(7):749–770.

2. Maron MS, Olivotto I, Zenovich AG, Link MS, Pandian NG, Kuvin JT, et al. Hypertrophic cardiomyopathy is predominantly a disease of left ventricular outflow tract obstruction. Circulation. 2006;114(21):2232–2239.

3. Ommen SR, Ho CY, Asif IM, Balaji S, Burke MA, Day SM, et al. 2024 AHA/ACC/AMSSM/HRS/PACES/SCMR Guideline for the Management of Hypertrophic Cardiomyopathy: A Report of the American Heart Association/American College of Cardiology Joint Committee on Clinical Practice Guidelines. J Am Coll Cardiol. 2024.

4. Arbelo E, Protonotarios A, Gimeno JR, Arbustini E, Barriales-Villa R, Basso C, et al. 2023 ESC Guidelines for the management of cardiomyopathies. Eur Heart J. 2023;44(37):3503–3626.

5. Olivotto I, Oreziak A, Barriales-Villa R, Abraham TP, Masri A, Garcia-Pavia P, et al. Mavacamten for treatment of symptomatic obstructive hypertrophic cardiomyopathy (EXPLORER-HCM): a randomised, double-blind, placebo-controlled, phase 3 trial. Lancet. 2020;396(10253):759–769.

6. Haraf R, Habib H, Masri A. The Revolution of Cardiac Myosin Inhibitors in Patients With Hypertrophic Cardiomyopathy. Can J Cardiol. 2024;40(5):800–819.

7. Garcia-Pavia P, Oręziak A, Masri A, Barriales-Villa R, Abraham TP, Owens AT, et al. Long-term effect of mavacamten in obstructive hypertrophic cardiomyopathy. Eur Heart J. 2024;45(47):5071–5083.

8. Desai MY, Seto D, Cheung M, Afsari S, Patel N, Bastien A, et al. Mavacamten: Real-World Experience From 22 Months of the Risk Evaluation and Mitigation Strategy (REMS) Program. Circ Heart Fail. 2025;18(1):e012441.

9. Desai MY, Wolski K, Owens A, Geske JB, Saberi S, Wang A, et al. Mavacamten in Patients With Hypertrophic Cardiomyopathy Referred for Septal Reduction: Week 128 Results from VALOR-HCM. Circulation. 2024.

10. Heinze G, Wallisch C, Dunkler D. Variable selection - A review and recommendations for the practicing statistician. Biom J. 2018;60(3):431–449.

11. Wallisch C, Dunkler D, Rauch G, de Bin R, Heinze G. Selection of variables for multivariable models: Opportunities and limitations in quantifying model stability by resampling. Stat Med. 2021;40(2):369–381.

12. Castrichini M, Alsidawi S, Geske JB, Newman DB, Arruda-Olson AM, Bos JM, et al. Incidence of newly recognized atrial fibrillation in patients with obstructive hypertrophic cardiomyopathy treated with Mavacamten. Heart Rhythm. 2024;21(10):2065–2067.

13. Desai MY, Hajj-Ali A, Rutkowski K, Ospina S, Gaballa A, Emery M, et al. Real-world experience with mavacamten in obstructive hypertrophic cardiomyopathy: Observations from a tertiary care center. Prog Cardiovasc Dis. 2024;86:62–68.

14. Reza N, Dubey A, Carattini T, Marzolf A, Hornsby N, de Feria A, et al. Real-World Experience and 36-Week Outcomes of Patients With Symptomatic Obstructive Hypertrophic Cardiomyopathy Treated With Mavacamten. JACC Heart Fail. 2024;12(6):1123–1125.

15. Boyle TA, Reza N, Hyman M, Supple G, See VY, Marzolf A, et al. Atrial Fibrillation in Patients Receiving Mavacamten for Obstructive Hypertrophic Cardiomyopathy: Real-World Incidence, Management, and Outcomes. JACC Clin Electrophysiol. 2025;11(2):411–413.

16. Masri A, Maksabedian Hernandez EJ, Wang Y, Gao W, Wu A, Han X. Real-World Adherence to Mavacamten in Patients With Obstructive Hypertrophic Cardiomyopathy in the United States. Circulation. 2023;148(Suppl_1):A12133–A12133.

17. Weissler-Snir A, Saberi S, Wong TC, Pantazis A, Owens A, Leunig A, et al. Atrial Fibrillation in Hypertrophic Cardiomyopathy. JACC Adv. 2024;3(9):101210.

18. Mizia-Stec K, Gimeno JR, Charron P, Elliott PM, Kaski JP, Maggioni AP, et al. Hypertrophic cardiomyopathy and atrial fibrillation: the Cardiomyopathy/Myocarditis Registry of the EURObservational Research Programme of the European Society of Cardiology. Open Heart. 2025;12(1).

19. Masri A, Kanj M, Thamilarasan M, Wazni O, Smedira NG, Lever HM, et al. Outcomes in hypertrophic cardiomyopathy patients with and without atrial fibrillation: a survival meta-analysis. Cardiovascular diagnosis and therapy. 2017;7(1):36.

20. Carrick RT, Maron MS, Adler A, Wessler B, Hoss S, Chan RH, et al. Development and Validation of a Clinical Predictive Model for Identifying Hypertrophic Cardiomyopathy Patients at Risk for Atrial Fibrillation: The HCM-AF Score. Circ Arrhythm Electrophysiol. 2021;14(6):e009796.

21. Hegde SM, Lester SJ, Solomon SD, Michels M, Elliott PM, Nagueh SF, et al. Effect of Mavacamten on Echocardiographic Features in Symptomatic Patients With Obstructive Hypertrophic Cardiomyopathy. J Am Coll Cardiol. 2021;78(25):2518–2532.

22. Ferrantini C, Scellini B, Vitale G, Pioner JM, Querceto S, Coppini R, et al. Mavacamten depresses human atrial contractility in the same EC50% range as human ventricle. Biophysical Journal. 2022;121(3):106a–107a.

